# A theoretical framework to quantify the tradeoff between individual and population benefits of expanded antibiotic use

**DOI:** 10.1101/2024.08.28.24312731

**Authors:** Cormac R. LaPrete, Sharia M. Ahmed, Damon J.A. Toth, Jody R. Reimer, Valerie M. Vaughn, Frederick R. Adler, Lindsay T. Keegan

**Author notes:** These authors contributed equally (Cormac R. LaPrete, Sharia M. Ahmed). These authors contributed equally (Frederick R. Adler, Lindsay T. Keegan). **Statements and Declarations Competing interests** The authors declare that they have no competing interests. **Author’s contributions** CRL, SMA, FRA and LTK developed and analyzed the model, CRL, SMA, DT, JRR, VMV, FRA and LTK drafted the manuscript. CRL, SMA, DT, JRR, VMV, FRA and LTK have all read and approved the manuscript.

## Abstract

The use of antibiotics during a disease outbreak presents a critical tradeoff between immediate treatment benefits to the individual and the long-term risk to the population. Typically, the extensive use of antibiotics has been thought to increase selective pressures, leading to resistance. This study explores scenarios where expanded antibiotic treatment can be advantageous for both individual and population health. We develop a mathematical framework to assess the impacts on outbreak dynamics of choosing to treat moderate infections not treated under current guidelines, focusing on cholera as a case study. We derive conditions under which treating moderate infections can sufficiently decrease transmission and reduce the total number of antibiotic doses administered. We identify two critical thresholds: the Outbreak Prevention Threshold (OPT), where expanded treatment reduces the reproductive number below 1 and halts transmission, and the Dose Utilization Threshold (DUT), where expanded treatment results in fewer total antibiotic doses used than under current guidelines. For cholera, we find that treating moderate infections can feasibly stop an outbreak when the untreated reproductive number is less than 1.424 and will result in fewer does used compared to current guidelines when the untreated reproductive number is less than 1.533. These findings demonstrate that conditions exist under which expanding treatment to include moderate infections can reduce disease spread and the selective pressure for antibiotic resistance. These findings extend to other pathogens and outbreak scenarios, suggesting potential targets for optimized treatment strategies that balance public health benefits and antibiotic stewardship.

## 1 Introduction

The global rise in antibiotic resistance poses a significant public health threat, leading the World Health Organization (WHO) to issue a warning that the world is “running out of antibiotics”.^[10;27]^ The emergence of antibiotic resistance adds complexity to the clinical challenge of ensuring that the right antibiotic is prescribed to the right patient at the right dose for the right duration, to maximize benefits and minimize harm.^[12;16]^ Antibiotic resistance necessitates balancing the potential benefits and risks of antibiotic use for individual patients alongside broader implications for public health. In some cases, antibiotic prescribing has clear benefits for patients that outweigh any public health concerns (e.g., life-threatening bacterial sepsis). In other cases, antibiotic prescribing has no benefits for patients (e.g., viral infections) while the potential individual harms are multi-fold: 1 in 5 patients experience side effects from antibiotics^[34;31]^; antibiotics have been shown to disrupt the gut microbiome, particularly in children where it can lead to conditions such as obesity^[21]^; and may increase an individual’s risk of developing future antibiotic-resistant infections^[25;23]^. For these situations, individual and public health benefits align and it is easy to strongly recommend antibiotic avoidance. The complexity arises in less clear-cut scenarios where the benefits to individuals are unclear. For example, in travelers’ diarrhea most patients recover without antibiotics; however, antibiotics can reduce the duration of symptoms, which, for some individuals, may be important^[8]^. In these cases, it is necessary to balance the individual benefit of reducing the duration of symptoms against the potential individual harms associated with broad-spectrum antibiotics, as well as public health harms including the development of antibiotic resistance. We propose that there may be scenarios under which prescribing antibiotics benefits public health; an idea absent from most antibiotic discourse (Figure 1). We suggest that prescribing antibiotics to an individual reduces disease transmission enough to reduce the overall size, duration, or existence of an outbreak. In this scenario, individual level harms are reduced as well the overall number of antibiotic doses used at the population level.

**Figure 1:**
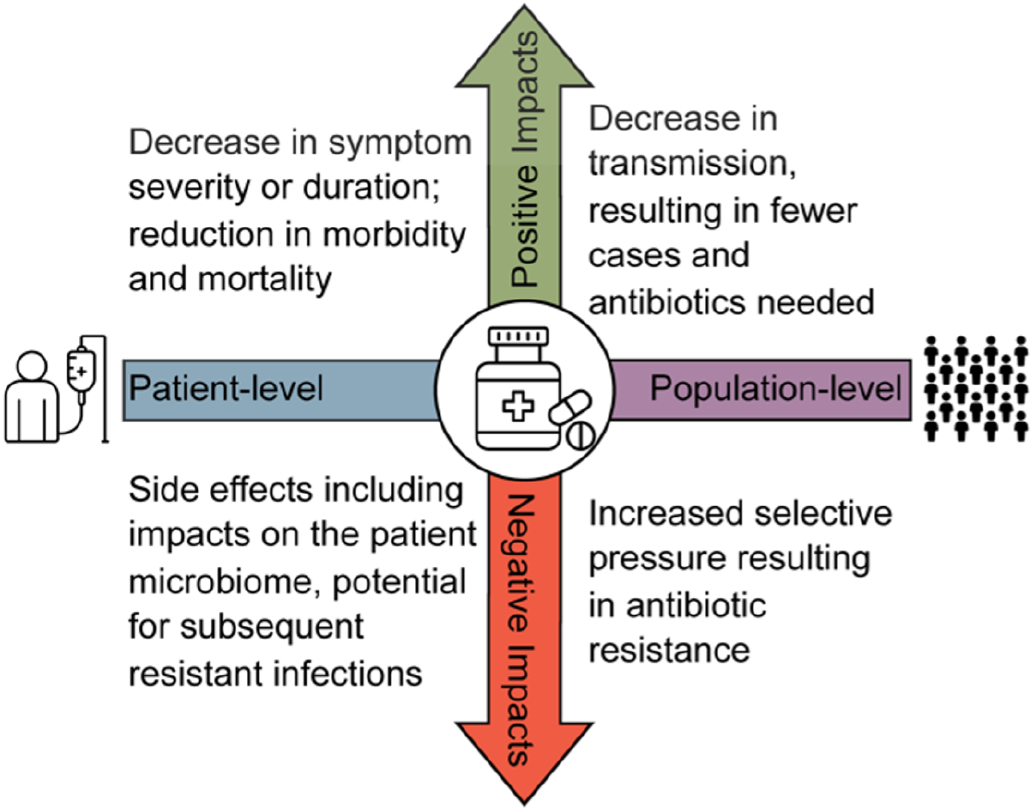
Schematic of individual and population-level harms and benefits of antibiotic use. The horizontal axis describes the individual (patient-level) and population-level impacts and the vertical axis describes the benefits (positive impacts) and harms (negative impacts) of antibiotic usage.

Here, we demonstrate a mechanism by which antibiotic use can offer population-level benefits through reduced transmission as a result of antibiotic treatment. That is, treating highly infectious individuals who may not require treatment to recover can reduce overall disease transmission, resulting in fewer total cases and/or fewer total antibiotic doses over the course of an outbreak. We explore this using cholera as a case study. Cholera, caused by the bacterium *Vibrio cholerae*, is a significant public health concern responsible for 1.3-4 million cases and 21,000-143,000 deaths annually worldwide^[2;4]^. Although most people infected with cholera do not develop symptoms, cholera can cause catastrophic diarrhea leading to potentially lethal dehydration. Since 2020, there has been a large increase in the number of cholera outbreaks, and these outbreaks have higher fatality rates than previously observed^[3]^.

The development of antibiotic resistance in cholera is a major concern that governs current antibiotic treatment recommendations^[1]^ which reserve antibiotics for patients with severe illness who are at highest risk of death without antibiotic treatment^[2]^. Patients with moderate illness generally experience self-limited symptoms that resolves with appropriate supportive care, and receive only a modest benefit from antibiotics primarily through reduction in symptom duration^[1;7;13;19;20]^. Although individuals with moderate infection may not individually be more infectious than severe cases, collectively, they can significantly contribute to onward transmission of the disease^[37]^. In these patients, antibiotic treatment can reduce shedding duration by up to 90%^[1;19]^. Without antibiotic treatment, an infected individual can shed cholera for up to 10 days^[2]^. Consequently, cholera provides an important opportunity to explore the tradeoffs between antibiotic utilization at both individual and population levels.

In this paper, we analytically solve for two thresholds to characterize these tradeoffs and use simulation to identify the conditions under which expanded antibiotic treatment to include moderate cholera infections presents a population-level benefit by reducing cholera transmission, outbreak persistence, and total antibiotic use.

## 2. Methods

### 2.1 Model

The authors declare that there is no associated data and code is available on GitHub (https://github.com/UT-IDDynamics/Cholera_Threshold).

To examine the tradeoffs between the individual- and population-level impacts of expanding antibiotic treatment, we analytically evaluate an extension of a *Susceptible-Exposed-Infected-Recovered* (SEIR) model (Figure 2). Here we explore the effectiveness of expanded antibiotic treatment at the population-level and assume that the individual-level benefit arises from reduction in symptom severity and duration. To do this, we compare the final size of the outbreak and the total number of doses given under different treatment scenarios, as well as the impact of these treatment decisions on outbreak emergence and spread.

**Figure 2:**
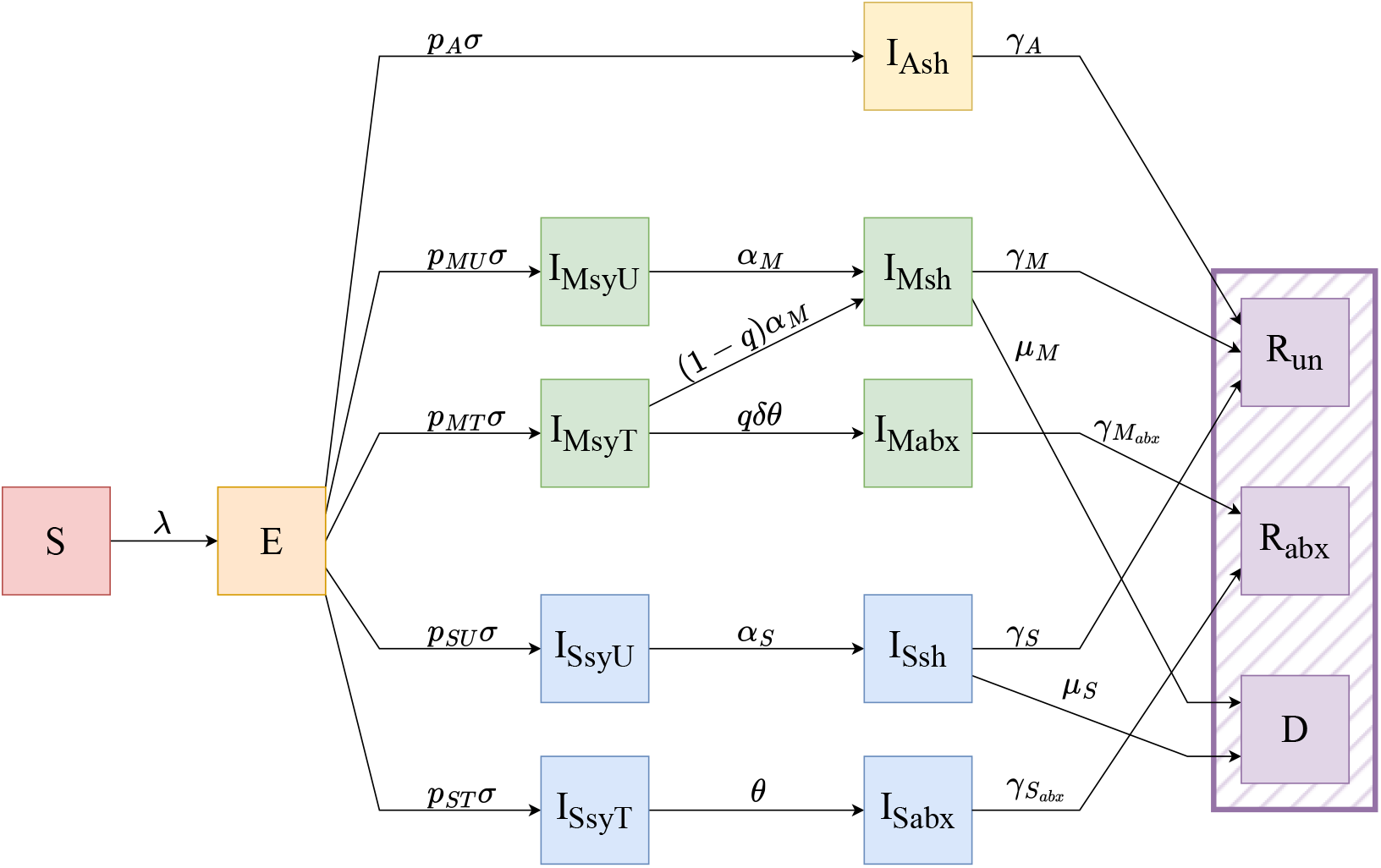
Compartmental model of cholera transmission dynamics. Each compartment represents a different epidemiological class. All individuals begin in the susceptible class (*S*), and become exposed (*E*) at rate *λ*. From *E*, individuals become infectious (*I*) and, based on their symptoms, follow one of five different paths (Supplement 4 further describes rates *p*_*A*_*σ, p*_*MU*_ *σ, p*_*MT*_ *σ, p*_*SU*_ *σ, p*_*ST*_ *σ*). Post-infection, individuals are removed in one of three ways (purple hatching: *R*_*un*_, *R*_*abx*_, *D*). Breaking down the transition from exposure to infectiousness, exposed individuals become asymptomatically infected (*I*_*Ash*_) at rate *p*_*A*_*σ*, are never eligible for treatment, and recover at rate *γ*_*A*_ to the recovered, untreated compartment (*R*_*un*_). Exposed individuals become infected with moderate symptoms (*I*_*MsyT*_ or *I*_*MsyU*_) at rate *p*_*MT*_ *σ* or *p*_*MU*_ *σ* depending on whether or not they seek treatment. Moderately infected individuals who do not seek treatment (*I*_*MsyU*_) recover from symptoms at rate *α*_*M*_ and continue to shed (*I*_*Msh*_) until they fully recover at rate *γ*_*M*_ to the recovered, untreated compartment (*R*_*un*_), or they die at rate *µ*_*M*_, moving to the dead compartment (*D*). Moderately infected individuals who seek treatment (*I*_*MsyT*_) may or may not receive treatment based on the model scenario. Treatment seeking patients with moderate infections receive treatment and recover from symptoms at rate *qδθ* or do not receive treatment and recover from symptoms at rate (1 − *q*)*α*_*M*_. Moderate infections who receive treatment continue to shed (*I*_*Mabx*_) for a shorter duration than those not receiving treatment, and fully recover at rate 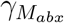 to the recovered, treated compartment (*R*_*abx*_). Treatment seeking patients with moderate infections who do not receive treatment recover from symptoms and continue to shed (*I*_*Msh*_) until they fully recover at rate *γ*_*M*_ to the recovered, untreated compartment (*R*_*un*_), or they die at rate *µ*_*M*_, moving to the dead compartment (*D*). Exposed individuals become severely infected (*I*_*SsyU*_ or *I*_*SsyT*_) at rate *p*_*SU*_ *σ* or *p*_*ST*_ *σ* depending on whether or not they seek treatment. Severely infected individuals who do not seek treatment (*I*_*SsyU*_) recover from symptoms at rate *α*_*S*_ and continue to shed (*I*_*Ssh*_) until they fully recover at rate *γ*_*S*_ to the recovered, untreated compartment (*R*_*un*_), or they die at rate *µ*_*S*_, moving to the dead compartment (*D*). All individuals with severe infections who seek treatment (*I*_*SsyT*_) receive treatment with antibiotics at rate *θ*. Those with severe infections who receive treatment continue to shed (*I*_*Sabx*_); however, they remain hospitalized until symptoms resolve and therefore do not contribute to transmission. They then fully recover at rate 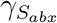 to the recovered, treated compartment (*R*_*abx*_). Full6model equations can be found in Supplement 4.

We model disease dynamics by dividing the population into the following compartments: Susceptible (*S*), Exposed (*E*), Infected (*I*), and Recovered (*R*); and further subdivide these compartments to capture the structure essential for our questions. To track disease outcomes more effectively, we differentiate the recovered class (indicated by a purple hashed box in Figure 2) by treatment status and death: Recovered untreated (*R*_*un*_), Recovered treated with antibiotics (*R*_*abx*_), or Dead (*D*).

We subdivide the infected compartment by symptom class, treatment seeking behavior, and shedding (Table 1). We split the infected compartment into three groupings by symptom classification, denoted with the following subscripts: Asymptomatic infections (_*A*_), Moderately symptomatic infections (which we call moderate infections) (_*M*_), and Severely symptomatic infections (which we call severe infections) (_*S*_). Asymptomatic infections do not cause noticeable symptoms but patients may still transmit. Moderate infections cause mild symptoms and patients may know that they are infected, but in the case of cholera, do not experience high fluid loss. Severe infections cause more serious symptoms, like heavy fluid loss, with an associated high risk of death if not treated promptly with re-hydration and/or antibiotics.

**Table 1:** Summary of infected states and their properties. This study focuses on states denoted with (*). The shedding column includes the associated term in the force of infection in Equation (1), illustrating the reductions in transmission relative to the shedding caused by severely symptomatic untreated infections (*β*).

| State | Symptoms | Treatment | Shedding |
| --- | --- | --- | --- |
| $I_{Ash}$ | Asymptomatic | Not seeking care, not treated | Full Asymptomatic ($\nu_A\beta$) |
| $I_{MsyU}$ | Moderate | Not seeking care, not treated | Full Moderate ($\nu_M\beta$) |
| $I_{MsyT}$ | Moderate | Seeking care, may receive antibiotic (*) | Full Moderate ($\nu_M\beta$) |
| $I_{Msh}$ | Moderate,<br>Post-symptom | May have sought care, but do not receive antibiotic (*) | Reduced Moderate ($\nu_M\nu_{sh}\beta$) |
| $I_{Mabx}$ | Moderate | Seen and treated with antibiotic | Minimal Moderate ($\nu_M\nu_{abx}\beta$) |
| $I_{SsyU}$ | Severe | Not seeking care, not treated | Full Severe ($\beta$) |
| $I_{SsyT}$ | Severe | Seeking care, will be treated with antibiotic | Full Severe ($\beta$) |
| $I_{Ssh}$ | Severe,<br>Post-symptom | Not seeking care, not treated | Reduced Severe ($\nu_{sh}\beta$) |
| $I_{Sabx}$ | Severe | Seen and treated with antibiotic | None |

We subdivide the moderate and severe infectious compartments based on whether or not they present to healthcare: seeking treatment (_*T*_) or not seeking treatment (untreated) (_*U*_). Because asymptomatic individuals do not experience symptoms, we assume that they never seek treatment and thus do not subdivide this compartment by treatment. We assume that those with severe infections are much more likely to seek treatment due to the dire nature of their symptoms, while only some individuals with moderate infections seek treatment.

Individuals shed at different rates based on symptom severity. Asymptomatic individuals shed less overall, whereas individuals who are untreated or have not yet received treatment shed at a higher rate (_*sy*_). Once symptoms resolve, moderate and severe infections maintain shedding for up to 10 days without antibiotic treatment (_*sh*_).^[2]^ We assume that all severely infected individuals receive antibiotic treatment (_*abx*_) and remain in a cholera treatment facility until shedding has resolved before they are discharged; thus we assume they do not contribute to transmission after treatment. On the other hand, because moderate infections do not require hospitalization alongside re-hydration, we assume they are able to transmit to others, although we assume that when treated (_*abx*_), moderate infections shed at a far lower rate per unit time than untreated severely infected individuals.

#### 2.1.1 Force of Infection

In this model, we define *β* as the rate of transmission by severely symptomatic infections and describe the transmission potential of all other infectious compartments relative to severely symptomatic infections. We do this because for cholera, untreated, severely symptomatic individuals shed the most per unit time and are thus the most infectious. As previously described, we assume that treatment with antibiotics, natural recovery, or infections with lower symptom severity all reduce transmissibility. We use different values of the parameter *ν* to represent the relative modification of infectiousness for individuals in these groups compared to untreated, severely symptomatic infections (Table 2). While each *ν* can take on any positive value dependent on the disease of interest, for cholera, we assume that *ν* ∈ [0, 1] to represent a proportional reduction in transmission. Individuals belonging to multiple groups multiply the values for each group.

**Table 2:** Model parameters and definitions, grouped by related parameters with subscripts to differentiate among symptom classifications, treatment, and shedding status. See Supplemental Table S1 (Supplement 4) for parameter values used in the simulations.

| Parameter | Description |
| --- | --- |
| $\lambda$ | force of infection |
| $\beta$ | transmission rate |
| $N$ | population size (use 1 to study proportion) |
| $p_A, p_{MU}, p_{MT},$<br>$p_{SU}, p_{ST}$ | proportion of exposed who become asymptomatic, moderately symptomatic, or severely symptomatic – these sum to 1 (Supplement 4) |
| $\sigma$ | rate of symptom development – inverse of latent period |
| $\nu_{sh}, \nu_A, \nu_M, \nu_{abx}$ | infectiousness relative to untreated severe symptoms of natural recovery, asymptomatic, moderate symptoms, and antibiotic treatment |
| $\gamma_A, \gamma_M, \gamma_{M_{abx}},$<br>$\gamma_S, \gamma_{S_{abx}}$ | recovery rate for asymptomatic, moderate, moderate treated with antibiotics, severe, severe treated with antibiotics |
| $\mu_M, \mu_S$ | death rate for moderate, severe |
| $\theta$ | treatment rate – inverse of time to treatment |
| $\delta$ | relative reduction of $\theta$ for moderate infection relative to severe |
| $\alpha_M, \alpha_S$ | rate at which symptoms resolve without antibiotic treatment (moderate, severe) |
| $q$ | moderate infection treatment effort |

The force of infection, *λ*, sums the contributions to transmission of all of the infected classes, except severely infected individuals who have received treatment 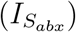, and is given by:

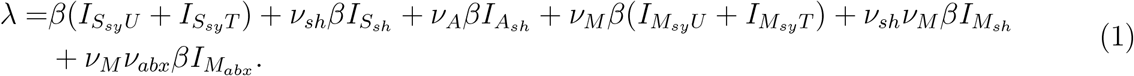

#### 2.1.2 Proportion of Moderate Infections Treated

To study the impact of treating moderate infections, we define the proportion of moderate infections that receive treatment as *M*_*abx*_. *M*_*abx*_ is the fraction of departure rates from 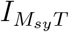 to the treatment states and depends on the treatment effort (*q*), recovery rate with antibiotic treatment (*α*_*M*_), and recovery rate without antibiotic treatment (*δθ*) (Table 2). *M*_*abx*_ is given by

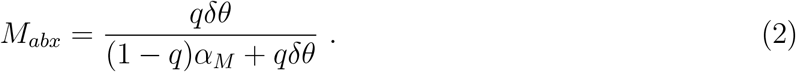

Because *q*, defined as the relative effort in treating moderate infections compared to treating severe infections, is difficult to measure in an outbreak setting, we rearrange for *q* in terms of the more easily measured *M*_*abx*_ to present model outputs. Rearranging, we find:

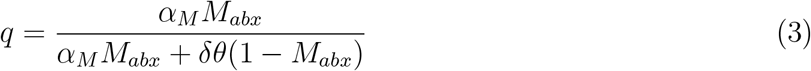

Both *q* and *M*_*abx*_ range from 0 to 1, where treating no moderate infections with antibiotics corresponds to *q* = *M*_*abx*_ = 0 (status quo), and treating all moderate infections that seek care corresponds to *q* = *M*_*abx*_ = 1.

#### 2.1.3 Cholera-specific assumptions

Several specific assumptions follow from our focus on cholera. First, the current guideline for treating cholera cases is to reserve antibiotics for severely symptomatic individuals who would be likely to die without such treatment.^[1]^ Under these guidelines, no moderately symptomatic infections are treated with antibiotics (*M*_*abx*_ = 0). Our study question explores the effects of increasing the proportion of moderates treated, *M*_*abx*_, between 0 and 1.

Next we assume that treatment with antibiotics reduces the transmission potential of an infected individual by reducing the duration of shedding in stool by 80-90%.^[1;7;13;19;20]^ Additionally, since symptoms severity is correlated with shedding volume, we assume lower severity infections are less transmissible.^[13;20]^ We use *ν*_*sh*_ (0.450) and *ν*_*abx*_ (0.500) to capture the reductions in shedding after symptoms resolve from natural recovery and from antibiotic treatment, respectively. For a comprehensive list of parameter values, see Table S1. These reductions apply to moderate and untreated severe infections, as we assume severe infections remain hospitalized until symptoms and shedding has resolved and therefore do not shed in the community. Finally, we denote the reduction in infectiousness of asymptomatic and moderate infections relative to severe infections by *ν*_*A*_ and *ν*_*M*_, respectively.

### 2.2 Reproductive Numbers

The reproductive number for a disease provides a population-level threshold for transmission. When the reproductive number is above 1, a disease will invade and persist and below 1 it will die out. We use this threshold condition to explore when expanded treatment may result in an outbreak dying out, by reducing the reproductive number below 1. Because we are interested in comparing the current treatment guidelines with expanded treatment guidelines, we calculate two reproductive numbers: the reproductive number under the current treatment guidelines, which we call, ℛ (q = 0), and the reproductive number under the expanded treatment guidelines, which we call, ℛ (q). Both are measures of transmission potential for an infectious disease, capturing the average number of secondary infections from a single infected individual.^[32]^

From equation (SI 1), ℛ (q) sums the contribution from each infection class (labelled in equation (4)) to the number of new infections in each generation, proportional to the total population (*N*)^[11]^. We find

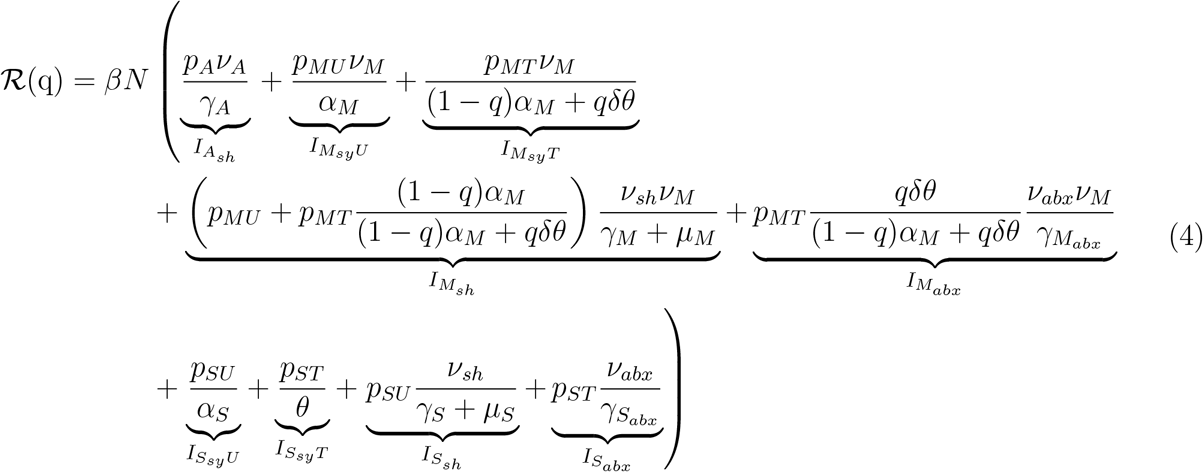

To find the reproductive number for the special case of no treatment, we substitute *q* = 0 into equation (4) and find ℛ (q = 0) to be

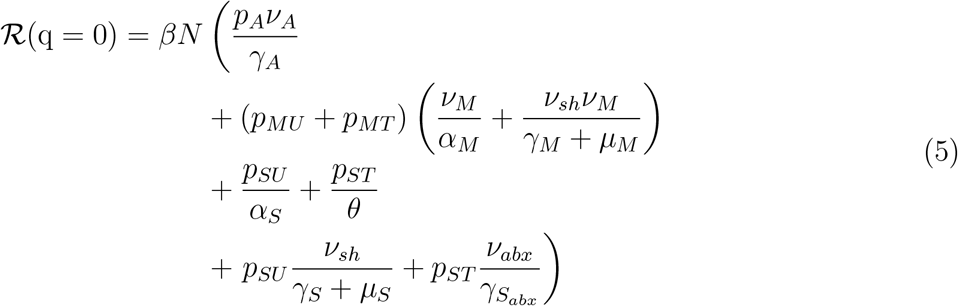

### 2.3 Outbreak Final Size

We compare the effectiveness of expanded antibiotic treatment on the final outbreak size, defined as the total proportion of infections over the course of the outbreak (i.e., the asymptotic state of the system).^[17;24]^ To do this, we separate the final size based on disease outcome and treatment status. We compare the number of untreated infections (*R*_*un*∞_), infections treated with antibiotics (*R*_*abx*∞_), and people who die (*D*_∞_) as well as the proportional final sizes *r*_*un*∞_ = *R*_*un*∞_*/N, r*_*abx*∞_ = *R*_*abx*∞_*/N*, and *d*_∞_ = *D*_∞_*/N*. These can be summed to find the overall final size of the outbreak, *r*_∞_ = *r*_*un*∞_ + *r*_*abx*∞_ + *d*_∞_.

Using the proportion of the population that remain susceptible at the completion of the outbreak (*s*_∞_), we derive equations (details in SI § 4) for the proportion of infections that were untreated (*r*_*un*∞_), the proportion of infections that were treated (*r*_*abx*∞_), and the proportion of infections that died (*d*_∞_).

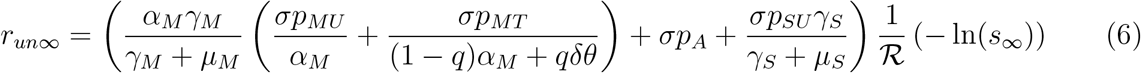

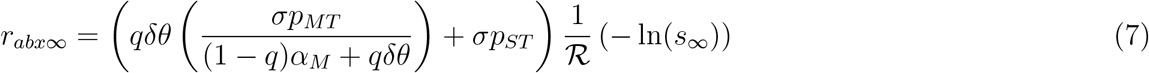

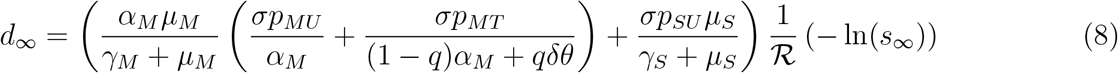

To understand the impact of treating moderate infections on the total proportions of infections and doses used in an outbreak, we numerically solve these final size equations (Equations 6 – 8) and vary the proportion of moderately infected individuals treated (*M*_*abx*_) from 0 to 1 and calculate the final size (*r*_∞_) using R Statistical Software^[30]^.

## 3 Results

Our analysis reveals two critical two-dimensional thresholds in the parameters ℛ (q = 0) and *M*_*abs*_ that summarize the relationship between antibiotic treatment and outbreak control. The first threshold occurs when expanded antibiotic treatment reduces the effective reproductive number (ℛ (q)) below 1, thereby halting the outbreak. We call this the Outbreak Prevention Threshold (OPT). The second threshold occurs when expanded treatment to include treating moderate infections reduces the number of antibiotic doses used over the course of the outbreak below that with the current treatment guidelines. We call this the Dose Utilization Threshold (DUT).

### 3.1 Outbreak Prevention Threshold (OPT)

To derive the OPT, we identify the conditions where expanded treatment reduces ℛ (q) below 1, stopping transmission. We define the OPT as the values of ℛ (q = 0) and *M*_*abx*_ for which an outbreak can be stopped with treatment of some proportion of moderate infections. For sufficiently large ℛ (q = 0), even treating all moderate infections will not reduce ℛ (q) below 1. We define ℛ_*opt*_ to be the maximum value of ℛ (q = 0) for which an outbreak can be feasibly stopped. To do this, we solved Equation (4) for ℛ (q) = 1, since ℛ_*opt*_ occurs when ℛ (*q* = 1) = 1. We then used Equation (2) to find values of *M*_*abx*_ from *q*.

With the parameter values for cholera, ℛ_opt_ = 1. 424. That means that for outbreaks with ℛ (q = 0) ≤ *ℛ*_opt_ = 1. 424, expanding antibiotic treatment to include moderate infections from the onset of the outbreak would prevent the outbreak from emerging and spreading. Because ℛ_*opt*_ describes the maximum value of ℛ (q) for which treating all moderate infections contains the outbreak, for lower values of ℛ (q), outbreak containment can be achieved by treating a smaller proportion of moderate infections.

The relationship between the proportion of moderate infections treated and the percent of the population infected (Figure 3A) is shown in Figure 3. Each curve that intersects the horizontal axis represents an ℛ (q = 0) value that can be reduced below 1 by treating some proportion of moderates. The proportion of moderates needed to contain the outbreak is given by the value where the curve intersects the horizontal axis. For curves that do not intersect the horizontal axis, even treating all moderate infections with antibiotics will not reduce ℛ (q) below 1.

**Figure 3:**
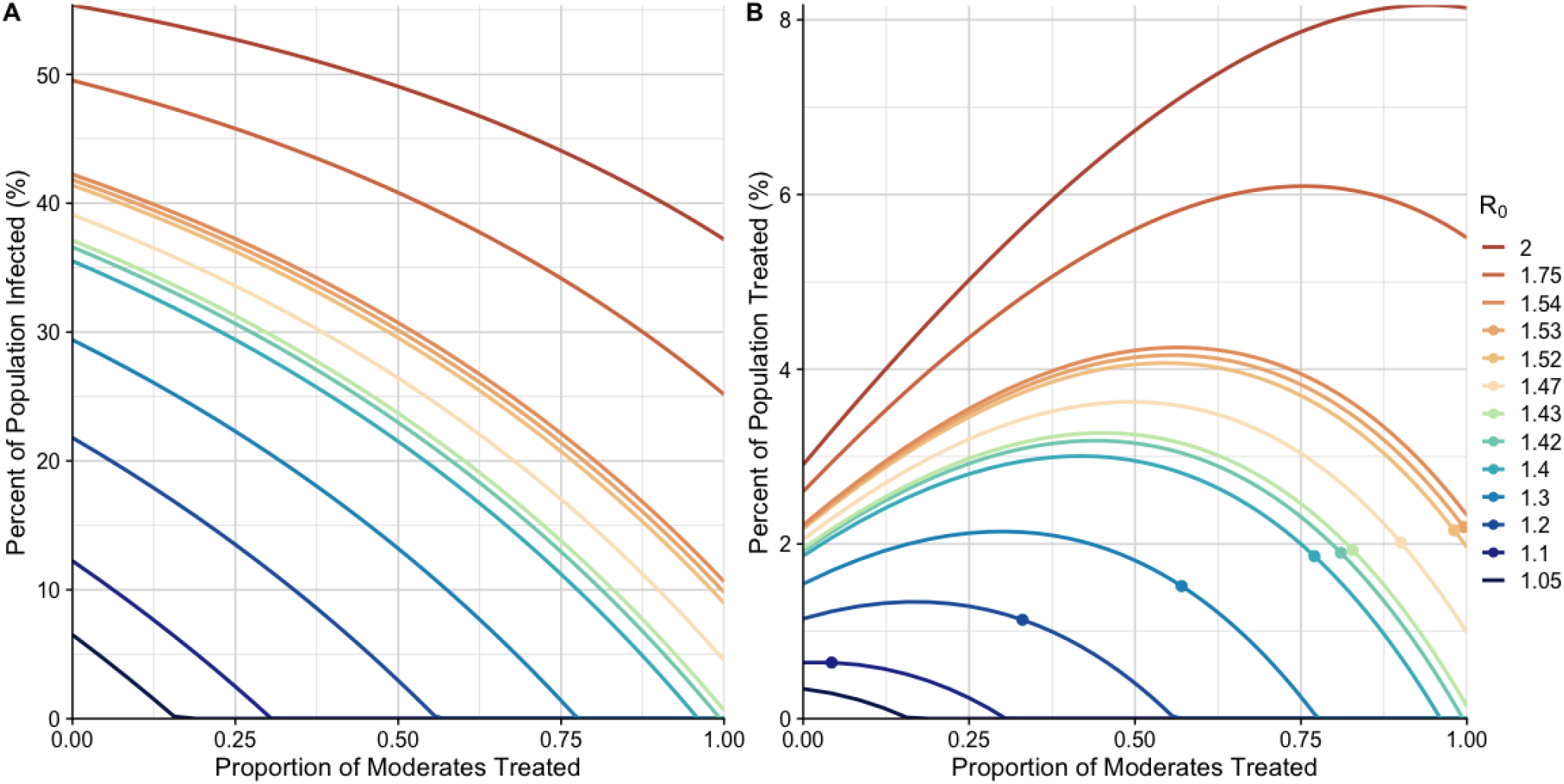
The final size (A) and proportion of total population treated (B) plotted against *M*_*abx*_ in a cholera outbreak for different values of ℛ (q = 0). Each curve corresponds to a different value of ℛ (q = 0), representing different outbreak scenarios of varying severity and transmissibility. (A) the percent of the population infected, regardless of severity of infection or treatment status, over an outbreak (*r*_∞_). (B) the percent of the population that receive antibiotics, including moderate and severe infections, over the course of an outbreak (*r*_*abx*∞_). Dots indicate the proportion of moderates that need to be treated to cross the Dose Utilization Threshold (DUT), for each value of ℛ (q = 0). Lines without points indicate that ℛ (q = 0) *> ℛ*_*dut*_.

**Figure 4:**
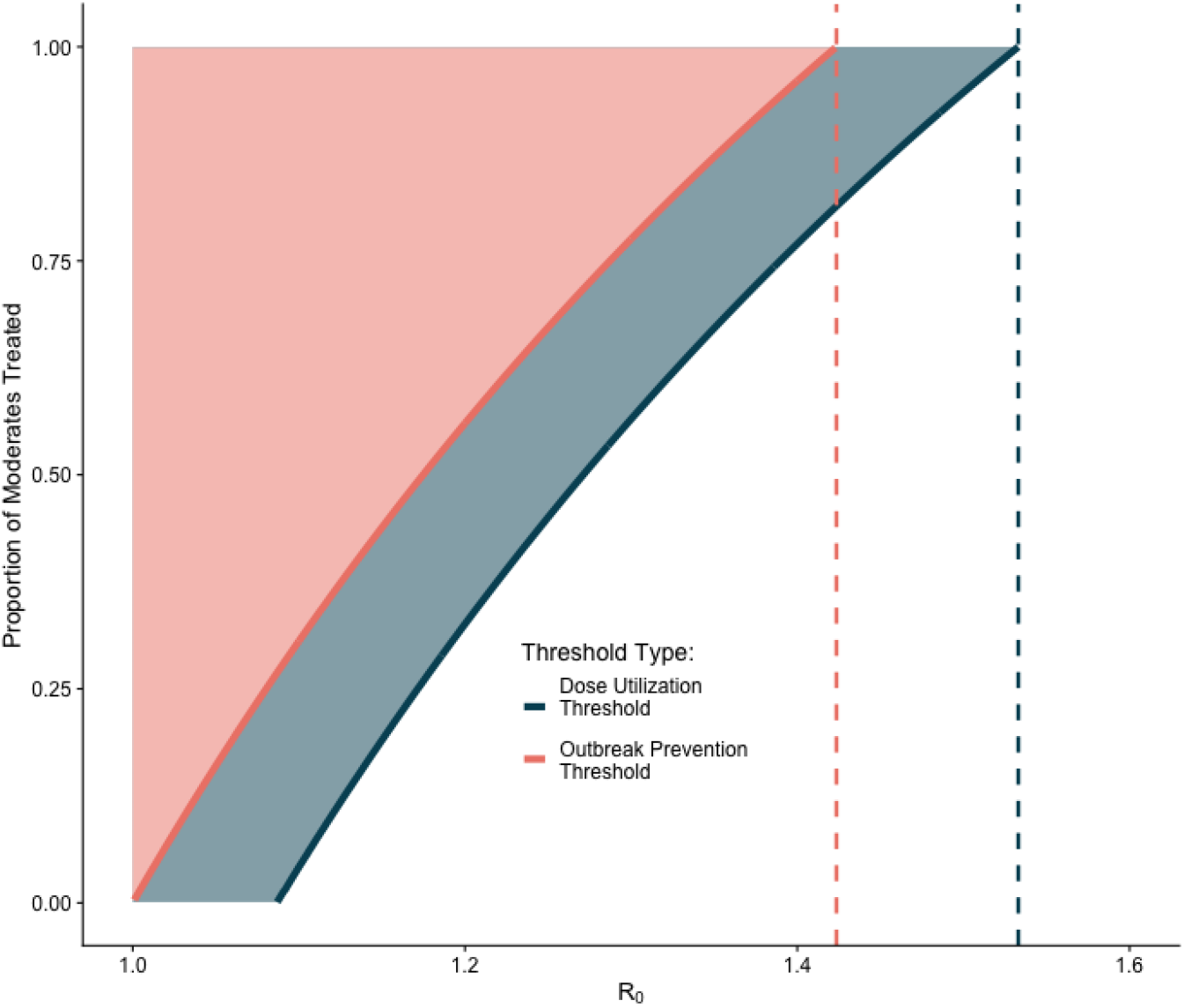
Plot of the threshold values for dose reduction (DUT) and outbreak prevention (OPT), by ℛ (q = 0), for each proportion of moderates treated (*M*_*abx*_). For a each value of ℛ (q = 0), the pink curve shows the OPT of ℛ (q = 0) and *M*_*abx*_, the shaded region above the curve shows all combinations of ℛ (q = 0) and *M*_*abx*_ in which treating moderate infections reduces the effective reproductive number, ℛ (q), below 1. The dotted red line indicates ℛ_*opt*_ for cholera. The blue curve shows the DUT of ℛ (q = 0) and *M*_*abx*_, the shaded region above the curve shows all combinations of of ℛ (q = 0) and *M*_*abx*_ in which fewer doses are given overall with expanded treatment. The dotted blue line indicates ℛ_*dut*_ for cholera. Outside of these regions, in the white space, there is no outcome improvement.

### 3.2 Dose Utilization Threshold (DUT)

We define the DUT as the values of ℛ (q = 0) and *M*_*abx*_ for which treating both moderate and severe cases with antibiotics results in fewer doses used over the course of the outbreak when compared to treating severe cases alone, where ℛ_*dut*_ is the maximum value of ℛ (q = 0) for which this occurs. To derive the DUT, we use the final size equations, Equations (6), (7), and (8). We identify the conditions under which expanding treatment can reduce the total number of doses used over the course of an outbreak. This can occur even if transmission is not completely halted and an outbreak still persists.

Mathematically, we solve for when *r*_*abx*∞_ (evaluated when *q* = 0) is greater than *r*_*abx*∞_ (evaluated when *q≠* 0). To find the threshold value for where this occurs, we set *r*_*abx*∞_(*q* = 0) = *r*_*abx*∞_(*q≠* 0), which results in the following equation

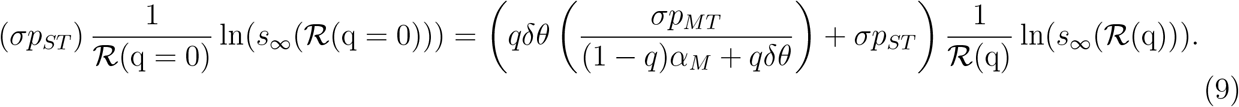

We numerically solve for ℛ_*dut*_, because this equation is not analytically tractable, as Equation (9) is an implicit function of ℛ (q = 0). For cholera we found ℛ_*dut*_ = 1.533. Like the OPT, for lower values of ℛ (q) a reduction in doses can be achieved by treating a smaller proportion of moderate infections. The relationship between the proportion of moderates treated and the number of doses of antibiotics used is shown in 4B.

While treating more cases (high *M*_*abx*_) will always reduce the number of cases in the outbreak (Figure 3A), the non-monotonic relationship between proportion of moderates treated and the number of doses of antibiotics used over the outbreak leads to an increase in the total number of doses used when when a small proportion of moderate infections are treated. However, as the proportion of moderate infections treated increases, the number of doses used decreases, and, for values of ℛ (q = 0) ≤ *ℛ*_*dut*_ = 1.533, drops below the number of doses used when treating no moderate infections (indicated by the points in Figure 3B). For outbreaks with very high values of *R*(q = 0), treating moderate infections substantially increases the total number of doses given as the reduction in transmission from treatment is insufficient to compensate for the large number of infected individuals (illustrated by *R*_0_ = 2 in Figure 3B).

## 4 Discussion

It has previously been thought that there is an unavoidable tradeoff between the utilization of antibiotics and the evolutionary risk of developing antibiotic resistance. Here we identify scenarios where antibiotic use creates a population-level benefit, thus benefiting both the individual and the population. The key mechanism is that antibiotics reduce the duration of shedding in treated individuals thereby reducing transmission and the total number of cases. Surprisingly, under certain circumstances, treating more patients with antibiotics can reduce the total number of doses used during an outbreak, thus reducing the selective pressures for the development of antibiotic resistance.

Here, we present a theoretical framework for finding the conditions under which treating moderate infections with antibiotics reduces the total number of doses used in an outbreak (which we term the Dose Utilization Threshold, or DUT) as well as when treating moderate infections completely stops an outbreak before it can take off (the Outbreak Prevention Threshold, or OPT). These two thresholds depend on two key measurable quantities, ℛ (q = 0) and *M*_*abx*_. When treating all moderate infections (*M*_*abx*_ = 1), we specify ℛ_*o*_*pt* and ℛ_*d*_*ut* as the maximum value of ℛ (q) for which these thresholds occur. When considering other values of *M*_*abx*_, the OPT and the DUT describe when these population-level benefits can be achieved by treating a smaller proportion of moderates for lower values of ℛ (q). However, if ℛ (q = 0) is larger than the DUT, the benefits of treating moderate infections is lost and the total number of doses given out over the course of the outbreak increases. This suggests that in some circumstances, it is better to not treat moderate infections if it is not certain that a threshold can be met due to concerns with public health infrastructure or compliance, for example.

Using cholera as case study, we found the conditions under which these thresholds occur. For cholera, we compute ℛ (q) ≤ *ℛ*_opt_ = 1. 424, below which treating moderate infections can reduce the effective reproductive number below 1, stopping the outbreak before it can spread. Similarly, when ℛ (q = 0) ≤ *ℛ*_*dut*_ = 1.533, treating moderate infections results in fewer doses used over the course of the outbreak than under current treatment guidelines. Because the range of reproductive numbers for cholera outbreaks is 1.1-2.7^[26;29]^, only outbreaks with low to intermediate values of ℛ (q = 0) can benefit from expanded antibiotic treatment. Further, since the relationship between the proportion of moderates treated and the number of doses used is non-monotonic, failure to treat a sufficient proportion of moderates treated may increase the number of doses used over the outbreak. The relationship between the proportion of moderates treated and the percent of the population infected is monotonical, any increase in the proportion of moderates treated reduces the percent of the population infected. Thus, careful evaluation of an outbreak setting, particularly with respect to community transmission (magnitude of ℛ (q = 0)), is essential prior expanding antibiotic treatment eligibility.

When ℛ (q = 0) ≤ *ℛ*_*dut*_, it is imperative to identify and treat sufficient moderate infections, particularly in the context of cholera, as many moderately infected individuals may not seek treatment. This is particularly true because the current treatment guidelines of reserving antibiotic treatment for severe infections may discourage treatment seeking behavior by those with more moderate cases. Therefore, even giving antibiotics to all moderate infections who present for treatment (as we do in our model, Table S1) may not be sufficient to achieve the population-level benefits iden-tified by our model. However, since individuals with moderate infections do not receive treatment under current guidelines, it is unknown how many would seek care if they qualified for antibiotic treatment under expanded access. This strategy may require proactively aiming to treat individuals who may not typically seek care and altering public messaging to encourage them to seek care. Further study is required to determine how treatment-seeking behavior may change should the policy change.

Our model relies on several simplifying assumptions, including that there is no heterogeneity in susceptibility. Our approach also relies on the assumption that severity is not transmitted; cases derived from moderate infections are equally likely to be severe as those derived from severe infections. We have also made a simplifying assumption by grouping infections into just three symptomatic classes. Because symptoms exist on a continuum, exploring a more granular division may help to further stratify the moderately symptomatic class by shedding and target the most infectious moderate infections for treatment. In our cholera case study, we used average reported parameter values. Additional sensitivity analysis could be used to determine which parameters have the greatest and most consistent influence on the threshold values and highlight them for future study. Our analysis evaluates only case counts and antibiotic use, neglecting other tradeoffs due to the economic costs of treatment^[35]^.

While we tailored our analysis to the specifics of cholera, this theoretical framework may be valuable for other pathogens. A similar framework has been considered in the context of Carbapenemresistant Enterobacterales (CRE) and decolonization in healthcare settings, where surveillance and testing strategies could be used to treat colonized individuals before they become infected, in parallel to the moderately symptomatic cholera patients considered here^[35]^. Similarly, for viruses such as influenza, this framework could aid in determining optimal strategies by balancing the tradeoff between testing and treatment. Here, factors such as cost-effectiveness^[33]^ and the development of resistance^[28]^ can play significant roles, as treatment strategies rely on individual vulnerability and susceptibility, as well as timing treatment before suspected symptom onset. This could be modeled to study scenarios in which it may be more beneficial to treat all cases as opposed to requiring a positive test for treatment. For HIV, particularly before the development of advanced antiretroviral therapies, healthcare providers had to balance side effects, patient compliance, and resistance concerns when trying to prevent HIV transmission prior to positive HIV test results.^[9;15]^ In such scenarios, our framework could help optimize the balance between side effects and compliance.

In conclusion, this study provides a new lens through which to view antibiotic use during infectious disease outbreaks, specifically highlighting the population-level benefits that can arise from expanding antibiotic treatment to include moderate infections. By shifting the emphasis from individual patient outcomes to broader population impacts, our findings challenge traditional approaches to antibiotic prescribing and propose a more strategic and context-sensitive framework for antibiotic stewardship.

## Data Availability

No data is produced in this study and all code produced is available online at: https://github.com/UT-IDDynamics/Cholera_Threshold.

## Acknowledgments

CRL, DJAT, FRA, LTK acknowledge funding from the Centers for Disease Control and Prevention (CDC) TRANSMIT: Training Research Acumen in Students Modeling Infectious Threats (Grant #1U01CK000675). SMA, DJAT, LTK acknowledge funding from the CDC Centers for Forecasting and Outbreak Analytics (CFA) (Grant #1NU38FT000009-01-00). VMV acknowledges funding from Agency for Healthcare Research and Quality (AHRQ) (Grant #5K08HS026530-06).

The authors thank Daniel Leung, Andrew Azman, and Iza Ciglenecki for helpful discussions about framing the research project.

## Supplementary Material

### Proportion of Exposed Becoming each Infected Type

Here we further describe the pathways for individuals becoming infected after exposure. The five proportions described in the main text (*p*_*A*_, *p*_*MT*_, *p*_*MU*_, *p*_*ST*_, *p*_*SU*_) can be broken down into more measurable decision-related values:

- *ϵ*_*A*_ = proportion of individuals exposed that are asymptomatic
- *ϵ*_*S*_ = proportion of all symptomatic individuals that have severe symptoms (1−*ϵ*_*S*_ = moderate symptoms)
- 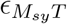 = proportion of moderately symptomatic individuals who seek treatment
- 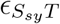 = proportion of severely symptomatic individuals who seek treatment

These allow the proportions to sum to 1 and the following equations describe the structure illustrated in Figure S1.

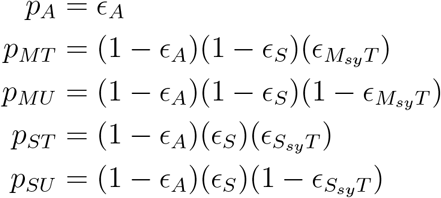

**Figure S1:**
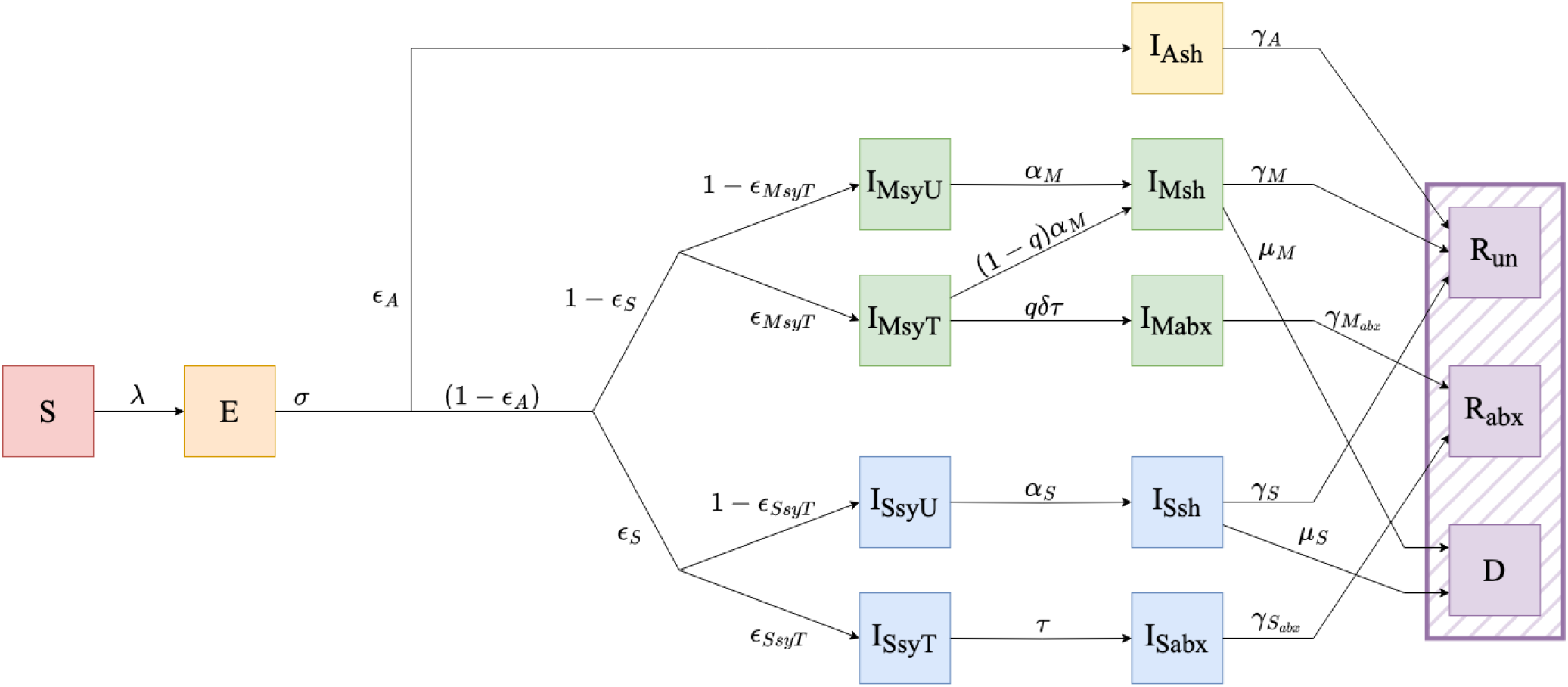
Model diagram with detailed parameterization of pathways between exposure and infection.

### Model Equations

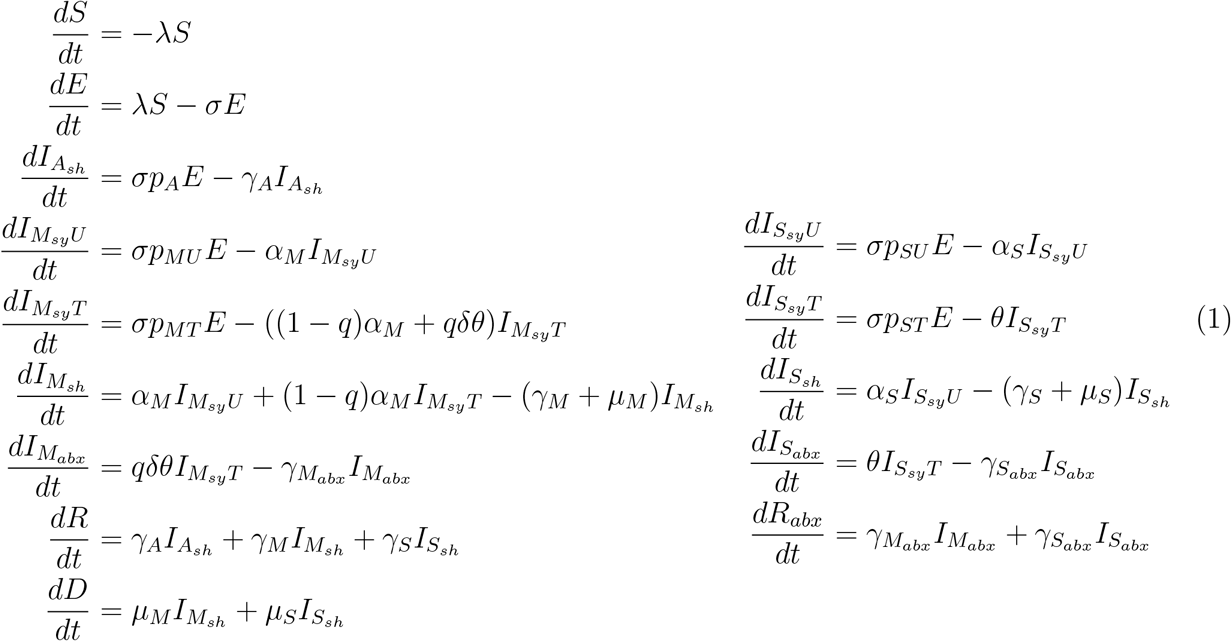

### Parameter values

**Table S1:** Model parameters values used in simulation with citations used to generate estimates. Note that ℛ_0_ is used to solve for *β* using equation (5).

| Parameter | Value | Citation |
| --- | --- | --- |
| $\mathcal{R}_0$ | 1.150 | [26;29] |
| $p_A$ | 0.750 | [6;14] |
| $p_{MU}$ | 0 | [6;14;37] |
| $p_{MT}$ | 0.175 | [6;14;37] |
| $p_{SU}$ | 0.023 | [6;14;37] |
| $p_{ST}$ | 0.052 | [6;14;37] |
| $\sigma$ | 0.695 | [5] |
| $\nu_{sh}$ | 0.450 | [13;20;36] |
| $\nu_A$ | 0.250 | see below |
| $\nu_M$ | 0.600 | see below |
| $\nu_{abx}$ | 0.500 | see below |
| $\gamma_A$ | 0.200 | [13;18;20;36] |
| $\gamma_M$ | 0.144 | [13;18;20;36] |
| $\gamma_{M_{abx}}$ | 1 | [13;18;20;36] |
| $\gamma_S$ | 0.060 | [13;18;20;36] |
| $\gamma_{S_{abx}}$ | 1 | [13;18;20;36] |
| $\mu_M$ | 0.01 | [2] |
| $\mu_S$ | 0.901 | [20] |
| $\theta$ | 2.664 | varies based on policies |
| $\delta$ | 0.500 | varies based on policies |
| $\alpha_M$ | 0.300 | [13;20] |
| $\alpha_S$ | 0.200 | [13;20] |

### Relative Infectiousness Modifiers

The values of *ν*_*A*_ and *ν*_*M*_ are not well characterized in the literature and would benefit from additional clinical research. For the purposes of this study, we find feasible values for these parameters (*ν*_*A*_ = 0.250, *ν*_*M*_ = 0.600) through a combination of literature review and expert elicitation. We know that severely dehydrated cholera patients have the longest duration of diarrhea symptoms (with and without antibiotic treatment), and the longest duration of culture positivity (with and without antibiotics)^[20]^. Counter-intuitively, the least dehydrated cholera patients with diarrhea had the second longest duration of diarrhea symptoms^[20]^. We assume that more days of diarrhea with less dehydration is indicative of lower volumes of stool. We further assume that lower volumes of stool equates to less bacteria released into the community, (*i*.*e*., less transmission). Finally, we assume that an absence of symptoms leads to better stool management (*e*.*g*., more likely to safely dispose of stool in covered pit latrine), further decreasing transmission for asymptomatic cholera patients compared to severely symptomatic cholera patients.

Similarly, *ν*_*abx*_ is not well characterized in the literature and would benefit from additional clinical research. For the purposes of this study, we find a feasible value for this parameter (*ν*_*abx*_ = 0.500) also through literature review and expert elicitation. It has been shown that among cholera patients treated with antibiotics, the proportion of patients testing positive by culture decreases rapidly over the first 2 days after treatment, with no culture positives individuals by day 4 ^[13]^. Similarly, it has been found that cholera patients were culture positive an average of 2.6 days after antibiotics^[20]^, whereas other studies documented an average of 1 day of stool culture positivity following antibiotic treatment^[36]^. While these estimates speak to the rate at which cholera patients treated with antibiotics recover, recovery is not a binary process where shedding is immediately halted. Therefore we opted for a conservative estimate of half transmission to encompass the time spent transitioning from the start of treatment to recovery.

### Final Size Equation Derivation

We rewrite here the relevant equations of the system and give them individual labels for reference.

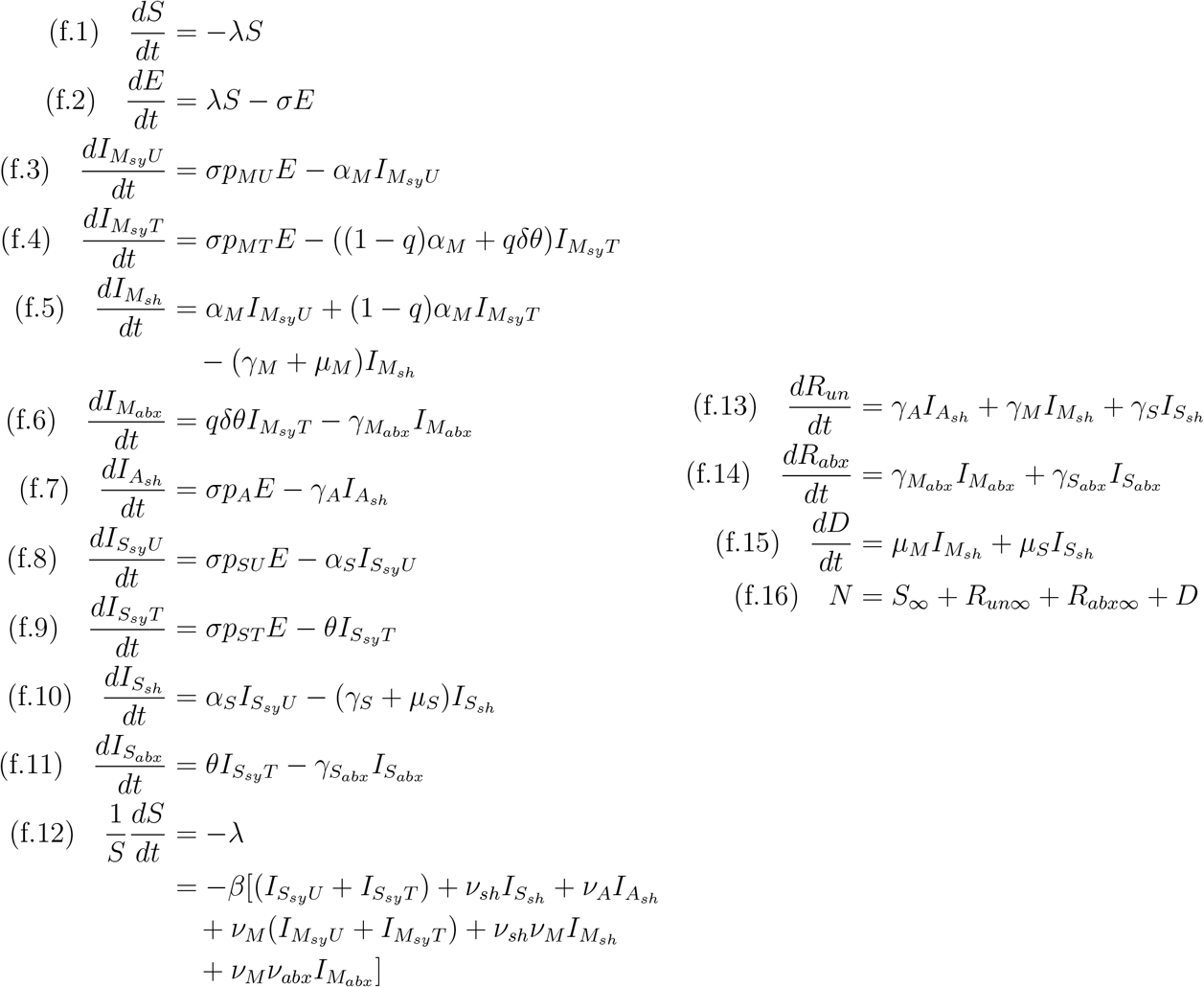

The following method avoids using (f.1) and (f.2) to avoid complications from the force of infection *λ*.

To find the final size, we integrate our equation over the time interval from 0 to ∞^[22]^. We start by setting the infected state equation integrals equal to 0 to simplify each equation in terms of ∫ *Edt*.

From setting the integral of (f.3) = 0:

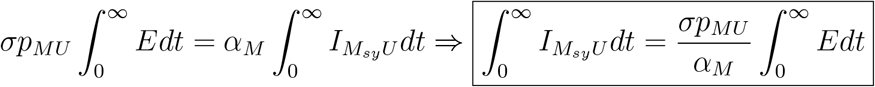

From setting the integral of (f.4) = 0:

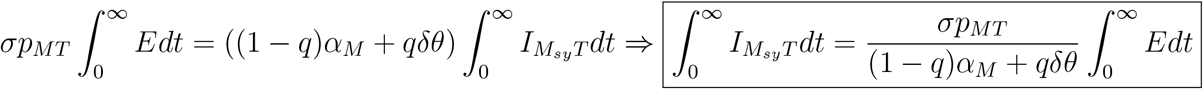

From setting the integral of (f.5) = 0:

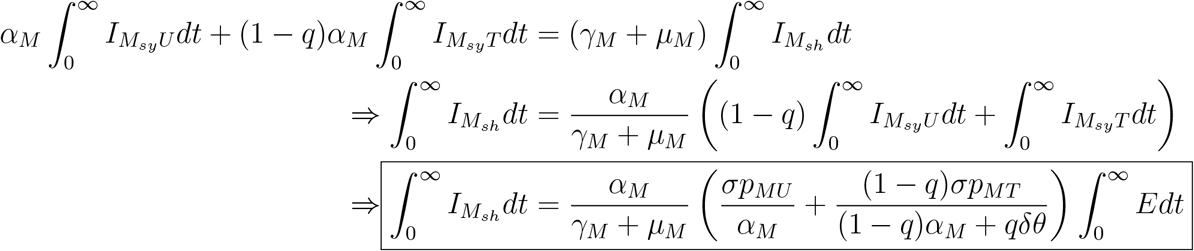

From setting the integral of (f.6) = 0:

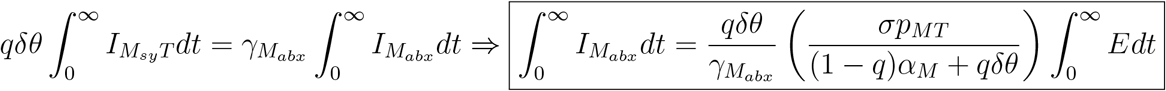

From setting the integral of (f.7) = 0:

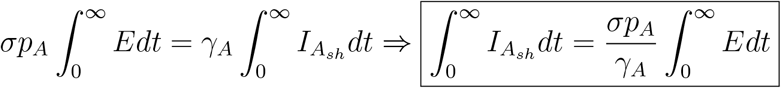

From setting the integral of (f.8) = 0:

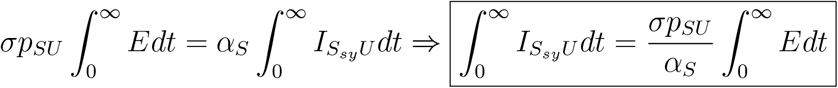

From setting the integral of (f.9) = 0:

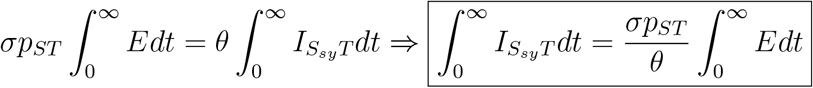

From setting the integral of (f.10) = 0:

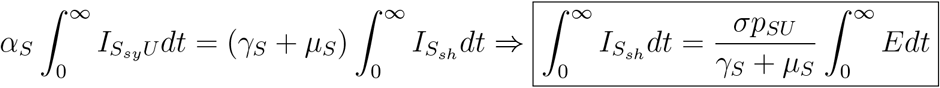

From setting the integral of (f.11) = 0:

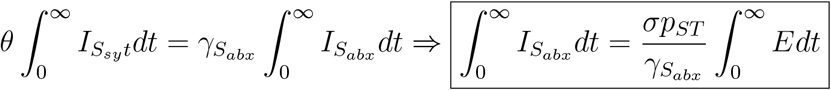

We then find the final sizes of the desired states in terms of f *Edt*.

From the integral of (f.13):

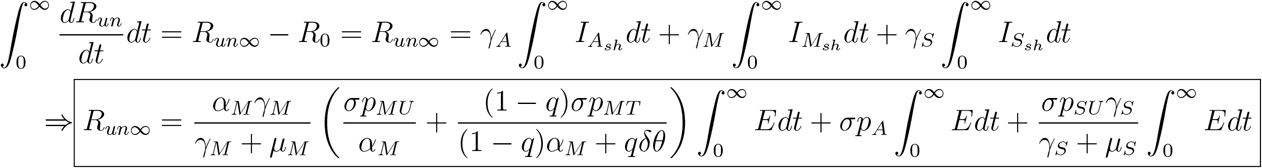

From the integral of (f.14):

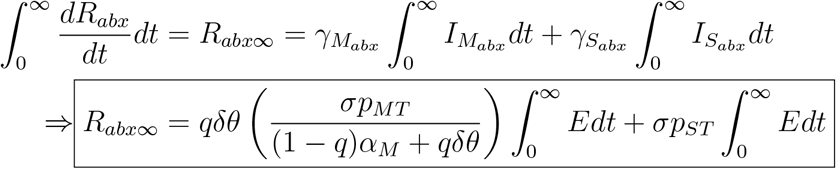

From the integral of (f.15):

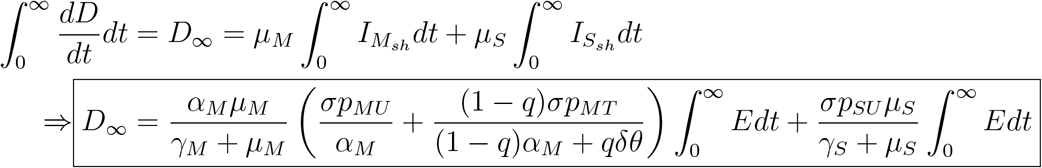

From here we want to find the ∫*Edt* to plug in above. We will use the trick of dividing both sides by *S*. From there we integrate and plug in the integrals from the infected states found above. (f.12) then integrates to solve for ∫*Edt*.

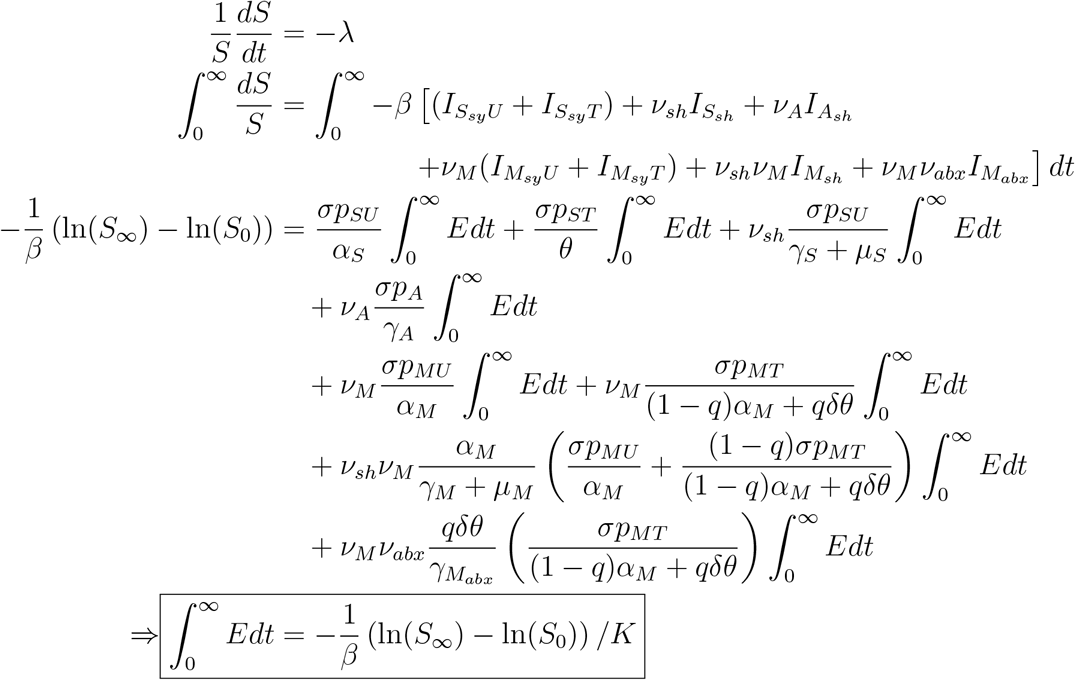

Where

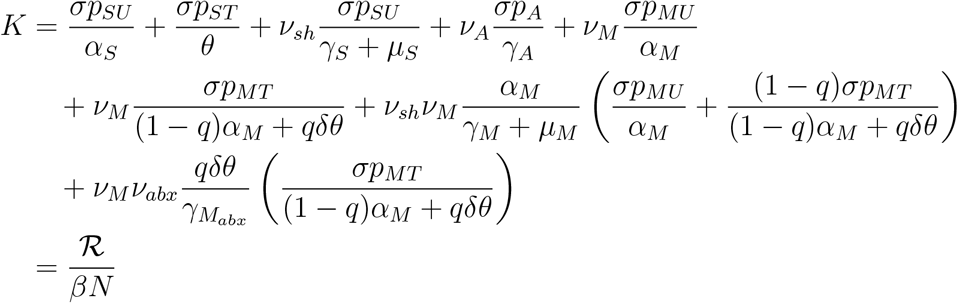

Therefore

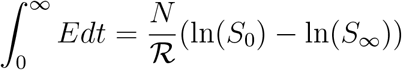

From here we use (f.16) to put all of the pieces together:

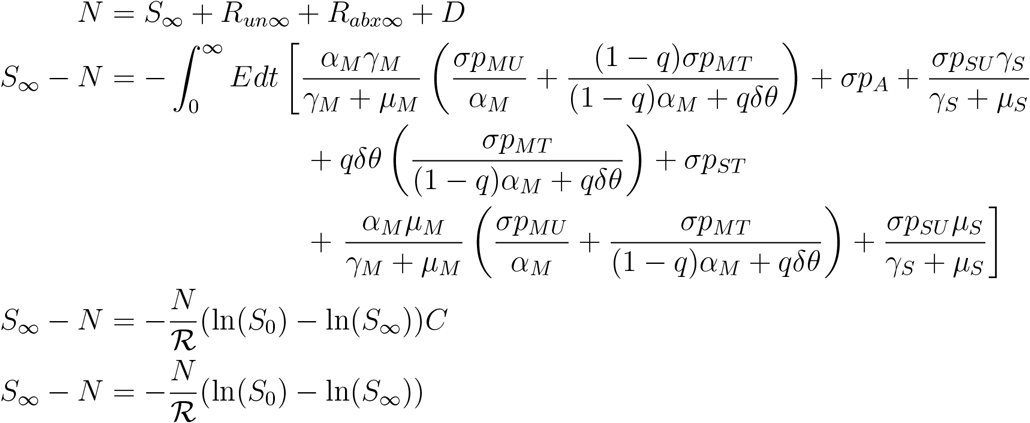

Where

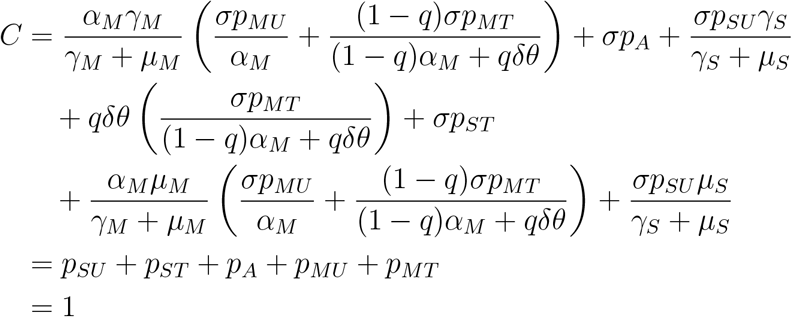

We continue to solve for *s*_∞_, where *s*_∞_ = *S*_∞_*/N*.

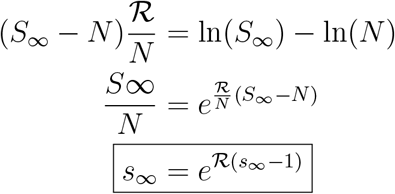

Importantly, this is the standard result for typical SIR and SEIR models as well.

So if we have

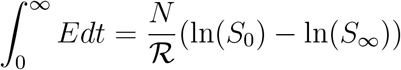

and

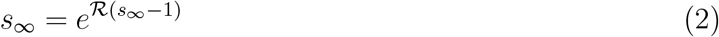

then we can get equations for the final sizes for *R*_*un*_, *R*_*abx*_, and *D* by plugging these into the equations for *R*_*un*∞_, *R*_*abx*∞_, and *D*_∞_ above after solving for *s*_∞_ numerically.

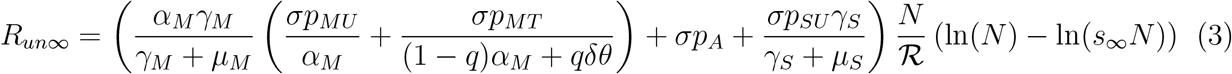

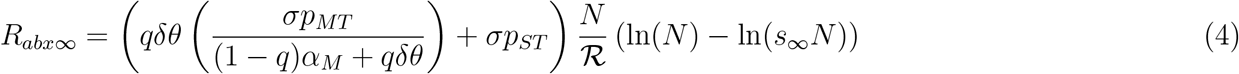

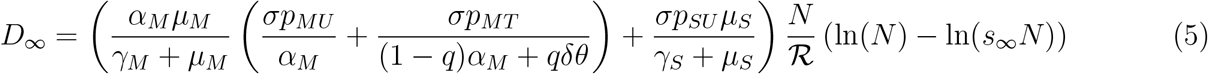

To study the proportions of individuals in each class, we divide out *N* and will call the proportional final sizes *r*_*un*∞_ = *R*_*un*∞_*/N, r*_*abx*∞_ = *R*_*abx*∞_*/N*, and *d*_∞_ = *D*_∞_*/N*.

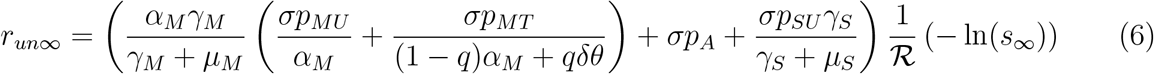

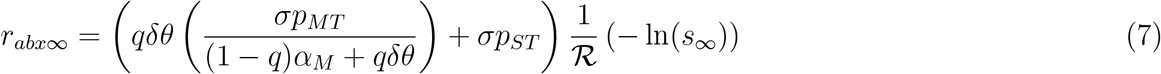

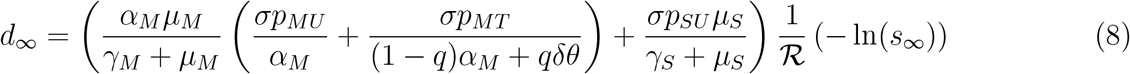

